# The Impact of Acoustic and Informational Noise on AI-Generated Clinical Summaries

**DOI:** 10.1101/2025.03.24.25324398

**Authors:** Thomas C. Draper, Jason Leake, Kathryn Lamb-Riddell, Timothy Cox, John McCormick, Stephen Trowell, Janice Kiely, Richard Luxton

## Abstract

**Objectives:** To investigate the effect of the introduction of environmental noise, microphone identity & position, and informational noise on the accuracy of a commercial Clinical AI Scribe (CAIS).

**Design:** Consultations on five medical conditions (memory loss, diarrhoea, headaches, skin rash, or prostate symptoms) were recorded. A ground-truth audio file was produced for a noise free consultation. This in turn produced a ground-truth clinical summary via the AI scribe, typically containing 30 facts. Different types of ‘noise’ at different levels were then investigated by (i) increasing the distance of the consultation from the microphone, and changing microphone type (ii) introducing background noise: heavy rain, baby crying, construction noise and a toddler at a range of levels between -10 to +5dB relative to the consultation audio and (iii) introducing informational noise: the medical consultations were modified to include additional conversations between the clinician and the patient to introduce irrelevant discussions (medical and non-medical, shorter and longer). The resulting 160 summaries were then compared with the corresponding ground-truth summaries and the errors introduced were classified and enumerated.

**Setting:** Simulated primary care setting.

**Participants:** The roles of the doctor and patient were played by actors, rotating between roles.

**Main outcome measures:** Outcomes measured were the quantification of the degradation of accuracy of the clinical summary, i.e. omissions, hallucinations and inclusions, relative to the ground-truth summary, following the introduction of noise of all types at different levels.

**Results:** Error rates and absolute error counts increased for all noise levels and noise types; the principal error being omissions. At 4.5 m from the consultation, for all microphones tested, all facts were omitted. At up to 2 m all microphones, except the laptop, only introduced a small number (<5) of omissions. For background noise, the type of noise was found to correlate with omissions – toddler and heavy rain were particularly deleterious, 7 and 15 omissions at 0 and +5dB respectively, i.e. when the noise loudness was comparable to or greater than that of the consultation. The CAIS was remarkably apt at rejecting additional informational noise. This was true for the addition of either short or long, medical or non-medical, information.

**Conclusions:** The CAIS was very good at capturing a doctor-patient consultation and producing accurate clinical summaries, however error rate (especially omissions) increased notably when acoustic noise was introduced. The system was better able to handle informational noise.

**Summary Box:** *What is already known on this topic:*

- The use of AI is rapidly increasing, with many commercial products available, both specific to clinical use and not.
- Commercial Clinical AI Scribes (generating clinical summaries from consultation audio) are being used in healthcare settings, sometimes with little guidance or knowledge of employers.
- These tools can operate well in perfect conditions, but there is limited data on nosier environments.

*What this study adds:*

- The commercial Clinical AI Scribe only introduced a small number of errors when introduced to informational noise, however large numbers of errors (especially omissions) were caused by acoustic noise (dependant on the particular sound type and volume), especially with low-quality poorly-located microphones.
- Users of Clinical AI Scribes must be particularly vigilant in checking the summary when the environment contains certain deleterious background noises, even at lower volumes, to ensure errors are not introduced to the patient records.

## Introduction

Recent studies have highlighted the transformative potential of AI in healthcare, e.g. see the reviews by Chustecki [1] and Al Kuwaiti [2]. The reviews highlight potential advantages of the technology together with risks and challenges, e.g. data security, regulatory issues and acceptance by patients and clinicians [3–5]. The most mature application is probably in the interpretation of diagnostic images, e.g. as produced by MRI, X-ray and CT scans [6,7]. The use of AI as a clinician decision support tool in general practice is also in development [8], as is the generation of discharge summaries and instructions for patients [9,10]. Recently, there has been a surge in the development of AI powered scribe technologies which transcribe clinician patient consultations into structured medical summaries [11]. There are now a significant number of such software packages available commercially. Reviews of some of these products are available via the KLAS website (https://engage.klasresearch.com/). Some of these products, together with databases of doctor patient encounters generated by these products, are also described in the bibliography of [11].

The AI medical scribe [12] adopts a two-step process (i) automatic speech recognition (ASR) produces a text transcript from the digital audio input which in turn is (ii) processed by a large language model (LLM) to produce a clinical summary. The summary is then input to the patient’s electronic health record (EHR). As there can be errors present in these summaries, it is currently mandated by commercial suppliers that the clinician should check the summary and correct for any errors prior to input to the patient’s EHR.

It has been proposed that the quality of the consultation is improved as the clinician does not need to make notes during the consultation and so is able to maintain improved eye contact with the patient [13]. The automated approach also has the potential to reduce the administrative burden on clinicians [14], thereby addressing the issue of burnout among clinicians [15]. It has also been reported [16] that the AI scribe can reduce the risk of errors which might be introduced by the clinician using manual input and increase the efficacy of EHR management.

Significant errors may be introduced by AI scribes; see for example Kernberg et al [17] who report significant errors for a ChatGPT-4 based scribe. Methods to determine and classify errors have recently been reviewed by Gebauer [11]. These include automated approaches, e.g. ROUGE and BERT [18–21] and also manual approaches [17] in which the clinical notes are compared with the original audio file, often by clinically trained personnel. A number of other metrics are also described, e.g. a modified Physician Documentation Quality Instrument-9 (PDQI-9) [22] and the Sheffield Assessment Instrument for Letters (SAIL) [23,24] approaches.

Despite the potential advantages, significant additional challenges exist in a real world clinical scenario in obtaining an accurate clinical summary of the consultation. The investigation of these challenges form the focus of the current paper: (i) firstly, the placement (and quality) of the microphone is critical to obtain an optimal audio input to the ASR; (ii) secondly, the clinical environment can be noisy with background noise present, e.g. from building work outside or the presence of a crying baby; (iii) and thirdly, informational noise may also be present during the consultation, e.g. a long and irrelevant diatribe about a hobby, or medical discussion which is not relevant to the consultation. These and other challenges have been described by Quiroz et al [25].

The effect of these additional challenges is investigated here experimentally. The approach that we have adopted is firstly to establish a ground-truth clinical summary with a good quality, noise-free audio file recording of a consultation, which is input to the Clinical AI Scribe. Noise, either acoustic (microphone or background) or informational (medical or non-medical), is then introduced at different levels to the audio file for the three challenges outlined above. The altered audio file is then presented to the Clinical AI Scribe to produce a modified clinical summary, which is then compared to the ground-truth summary to establish the additional errors introduced by the presence of the introduced noise.

## Methods

### Study Design

All medical consultations used, recorded, or referred to in this study were performed using actors and simulated medical conditions. For full protocol details, see the Protocol file in the supplementary information.

This research was conducted in a simulated primary care setting at the NHS South West Centre of Digital Excellence (CoDE). This provided a simulated GP surgery, allowing for increased realism in the positioning of persons and equipment, background noise, and acoustic profile. A controlled experimental framework was implemented to assess the impact of microphone cost (RRP £25–£120) and placement, background noise, and varying informational noise (both medical and non-medical) on the accuracy of a widely-used commercial Clinical AI Scribe. A repeated-measures approach was adopted, enabling systematic variation of these factors while maintaining as consistent an input to the AI as possible. Specifically, simulated consultations (typical 10 min in length) were recorded and presented to the clinical AI scribe, which generated a clinical summary (containing typically 25–30 points of information). The original audio files were then modified (for example, by including construction noise or a prolonged conversation about tennis) and re-presented to the same Clinical AI Scribe, which generated a new clinical summary, which could be compared to the original summary. These processes are displayed in figure 1.

**Figure 1:**
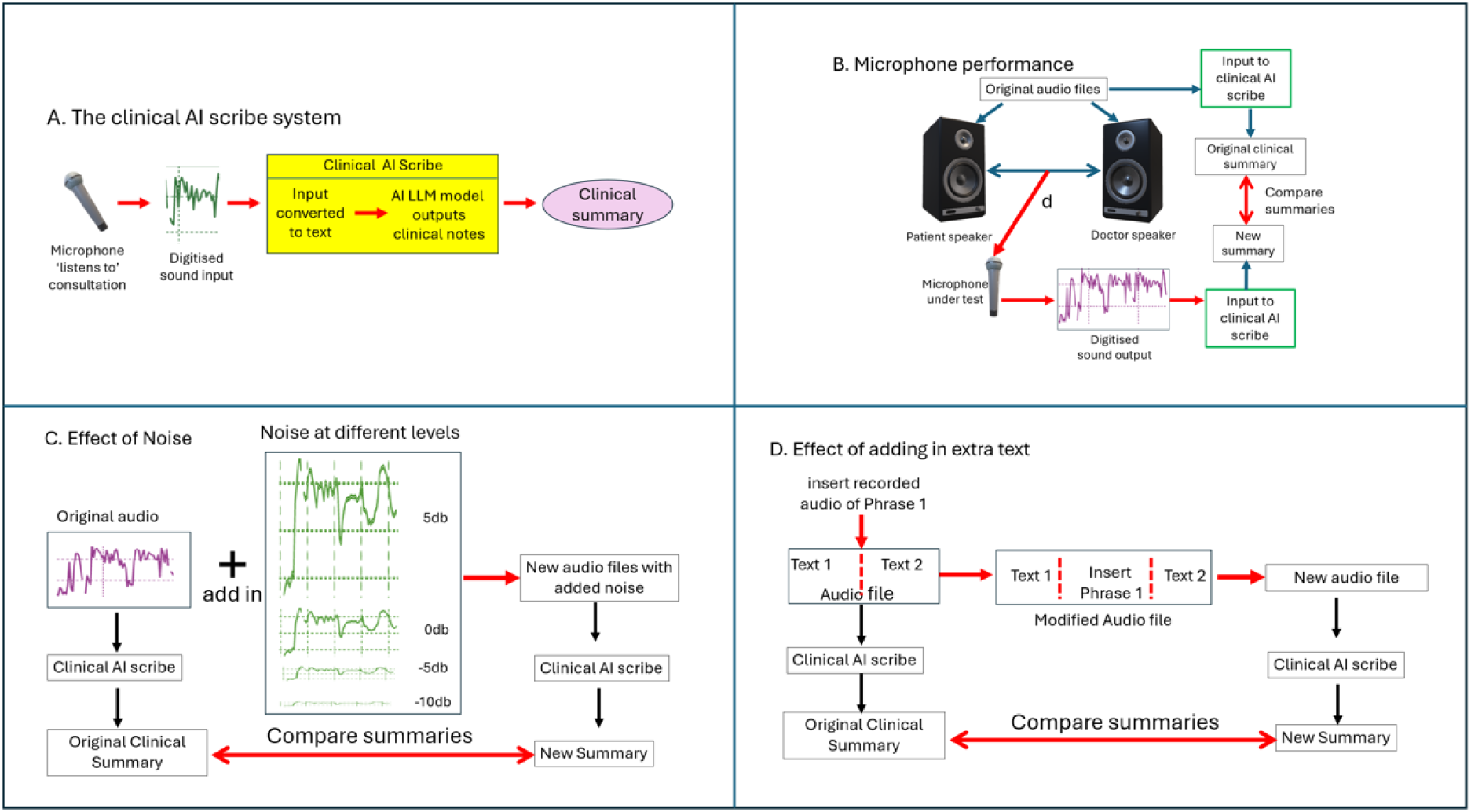
A graphical representation of the study design. (A) The general process for using a clinical AI scribe, from the audio being detected by the microphone through the CAIS, resulting in a clinical summary. (B) The process used to investigate microphones and their positioning. Different microphones, at different positions, were used to record a consultation, then all of the resulting summaries were compared to the original ‘clean’ summary. (C) The process used to investigate different background noises. To the original audio was added various noises at various volumes, which were then presented to the CAIS, the resulting summaries from which were then compared to the original ‘clean’ summary. (D) The process used to investigate varying informational noise. Medical or non-medical monologues or conversations were inserted into the original audio file, which are then presented to the CAIS, the resulting summaries from which were then compared to the original ‘clean’ summary.

When comparing clinical summaries, three measures are used:

- Omissions: These are points of information which are present in the original clinical summary, but not present in the compared summary.
- Hallucinations: These are points of false information which are not present in the original clinical summary, but are present in the compared summary.
- Inclusions: These are points of truthful information which are not present in the original clinical summary, but are present in the compared summary.

This is similar to the approach of Kernberg et al. [17], but with the addition of inclusions due to the inter-summary nature of our comparison.

### Patient and Public Involvement Statement

A Public/Patient Group (Citizens Reference Group) has been set up for the NHS Healthcare Centre of Digital Excellence. They were consulted in the design phase of the study and were provided with early-stage results to which they gave feedback to further inform the study.

## Results

### Impact of Microphone Placement (Acoustic Noise)

The distance between the participants of the consultation and the recording microphone is a key factor in ensuring high quality output from the clinical AI. Four microphone positions were looked at: control, 0.5 m, 2.0 m, and 4.5 m, chosen to simulate a seated conversation, small movement (e.g. to do test), and larger movement (e.g. moving to couch or behind examination screen). As can be seen in figure 2, the total points of information reduces as the distance is increased. This is expected because as the distance is increased the intensity of the sound signal at the microphone decreases. This might be expected to follow an inverse square law, *I* ∝ *d*^−2^ where *I* is the audio intensity and *d* is the distance. The decrease in the intensity results in a reduction in the bit depth of the recorded speech, whilst the bit depth of the background noise remains constant, ultimately resulting in a sharp decrease in the signal-to-noise ratio, and so the accuracy of the text produced by the ASR.

**Figure 2:**
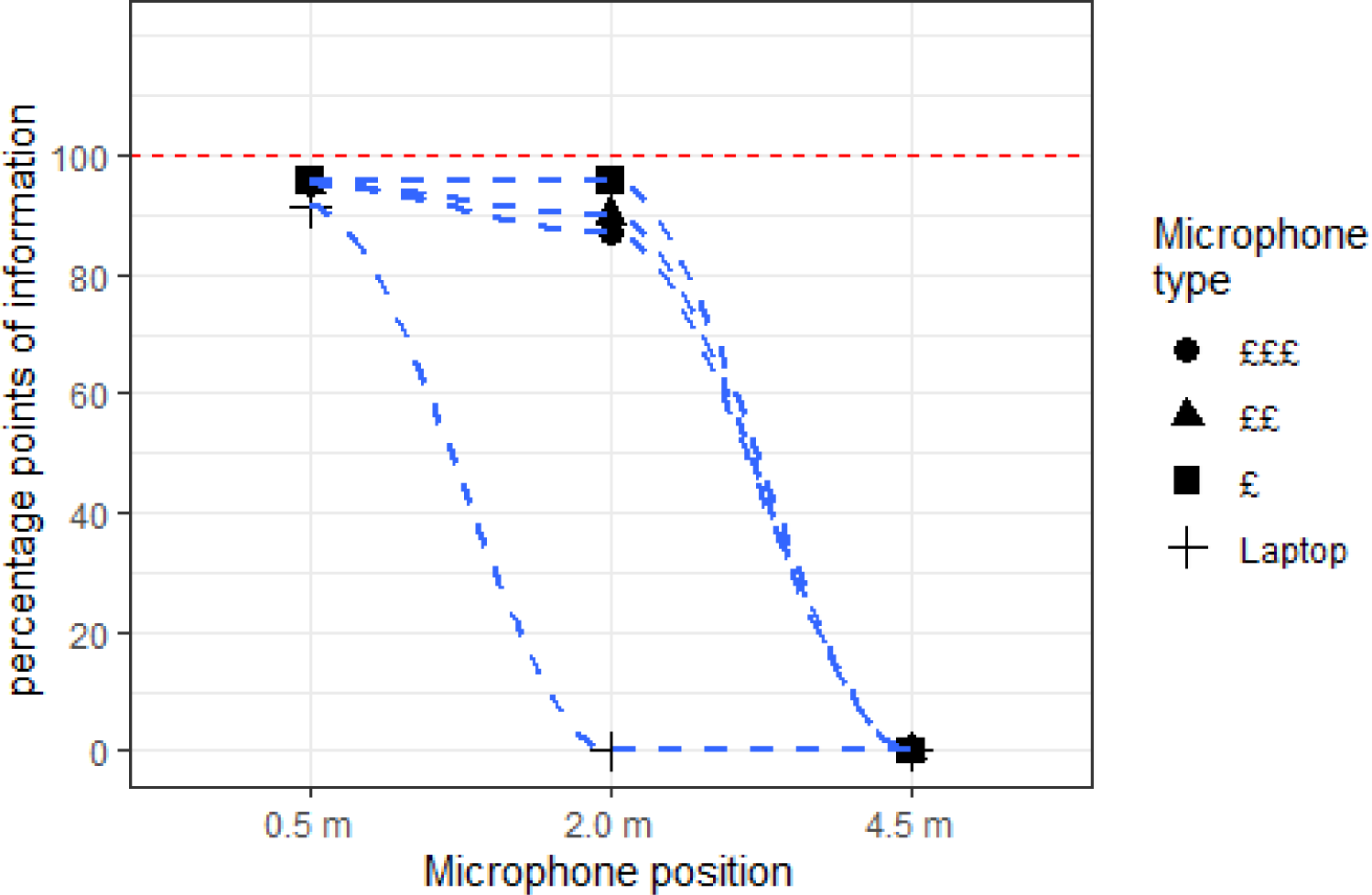
The median points of information, as a percentage, included in a clinical summary produced by an AI with different microphones (having different costs, see Protocol supplementary information), as a function of distance and relative to the original summary, for all five medical conditions combined. The blue dashed lines are to guide the eye.

This decrease in the points of information as the distance increases correlates with the increase in the numbers of errors, as shown in figure 3. The number of hallucinations and inclusions remains relatively constant across the distances, though the number of omissions rapidly increases, and in many cases the number of omissions equates to the original number of points of information (thus the complete loss of information at 4.5 m, shown in figure 2). These omissions occur because of a failure of the speech-to-text (ASR) process, which is unable to accurately recognise the English speech. A lack of inclusions is anticipated, as the introduction of additional truthful information is unlikely when the original information has already been compromised. Of note however, is the small but visible peak in hallucinations at the 2m position. This is primarily caused by the cheapest of the tested microphones, which had a surprising tendency to cause hallucinations at this distance. At 0.5 m, the signal is sufficiently robust to permit operation with minimal errors, whereas at 4.5 m the signal is so weak that the AI is unable to continue processing; however, at 2 m, an “anti-Goldilocks zone” is observed. In this region the LLM assumes a sufficient quantity and quality of information to proceed, but is overconfident.

**Figure 3.**
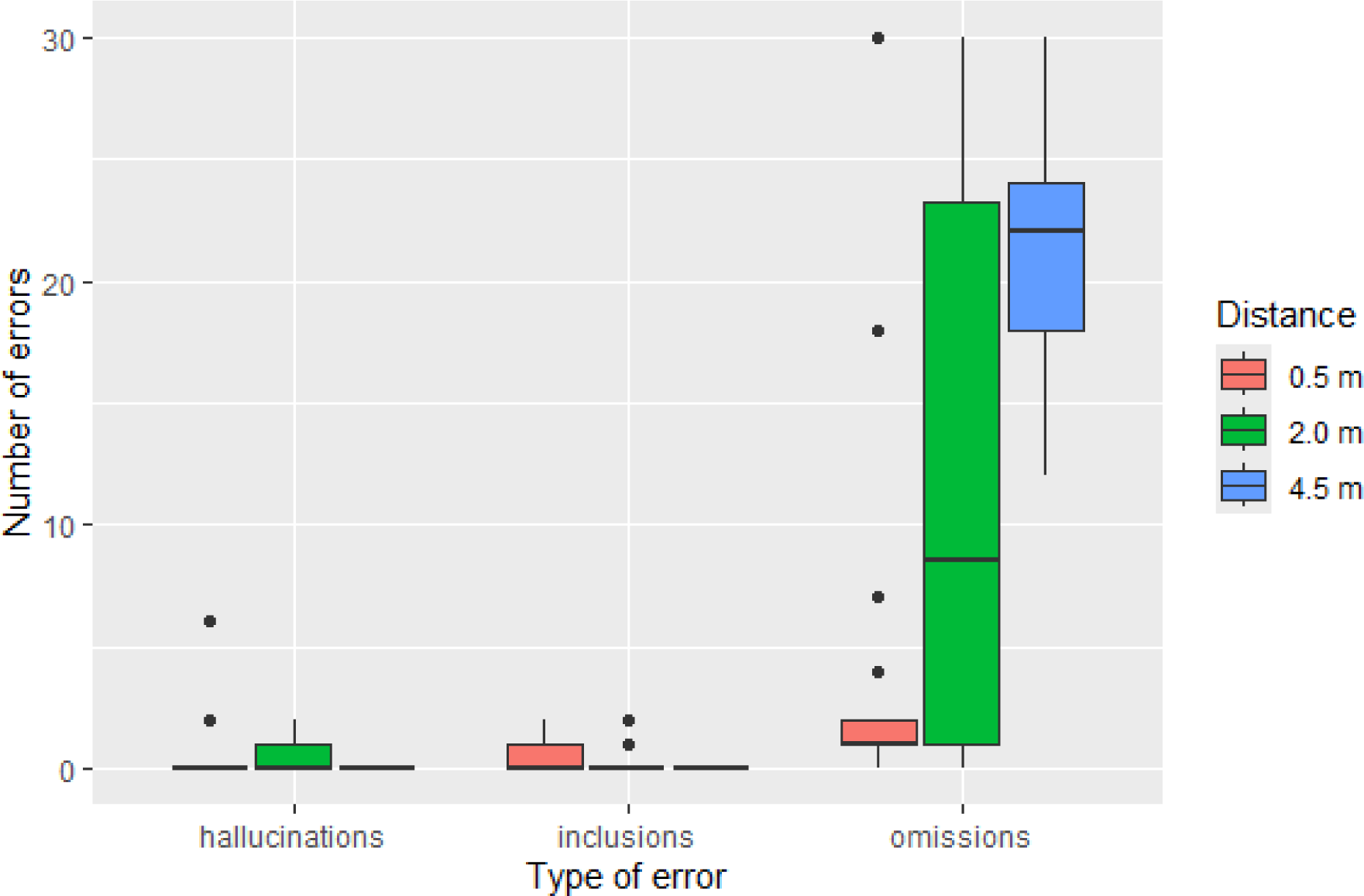
The number and types of errors at different microphone positions, d: 0.5 m, 2.0 m, and 4.5 m from the midpoint of the source, for all microphones and medical conditions combined.

It is interesting to note that, in most cases, the cost/brand of the microphone has little impact on the performance of the clinical AI, with all but one sharply dropping off after the distance is increased beyond 2 m. The sole exception to this is the laptop integrated microphone, which deteriorated at a much greater rate. This can also be seen in figure S1 in the supplementary information.

### Impact of Background (Acoustic) Noise

In a real-world primary care setting, there could be significant confusing sounds present, e.g. generated by those present in the consultation and also outside the consulting room. Four different typical background noises were investigated at varying volumes: baby crying, construction noise, heavy rain, and toddler chatter (for source details, see supplementary information). All four resulted in the introduction of errors, with the number of errors typically increasing as the volume of the background noise, relative to the consultation sound level, was increased. This increase can be seen in figures 4 and S2 (supplementary information). The volumes used were 0 dB (approximately the same volume as the consultation), -10 dB (about half the perceived loudness, or ∼10% of the sound pressure intensity), -5 dB (∼30% of the sound pressure level, i.e. noticeably quieter), and +5 dB (∼3x sound pressure level, i.e. noticeably louder). For reference, the background noise could be clearly discerned at -10 dB, whilst at +5 dB the speech was still comprehensible, though the consultation would be unlikely to proceed until the noise had abated. Moderate rainfall producing noise levels around -5 dB in an office is within the realm of possibility; however, rainfall sufficient to reach +5 dB is less likely. In contrast, internal noise contributions (e.g. from a toddler or baby) have a higher probability of reaching the +5 dB threshold.

**Figure 4:**
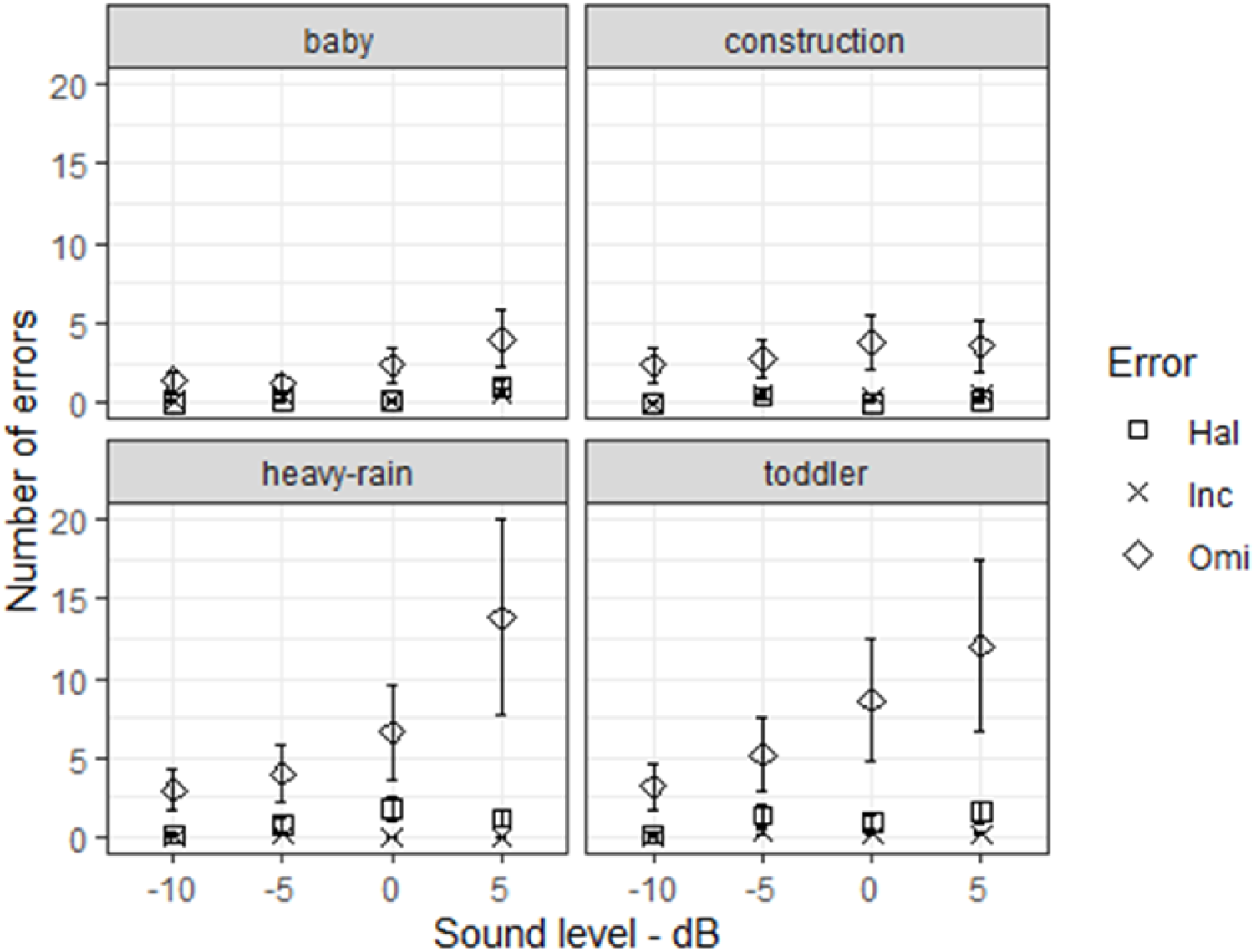
The number (and type) of errors introduced when different volumes of the four background noises are introduced. Four different sources of background noise are presented, with results for all medical scenarios combined. Hal: Hallucination, Inc: Inclusions, Omi: Omission. Values shown are the mean, and errors bars are standard error.

When comparing directly between volumes, across all background noise sources, there was no statistically significant increase in error rate between the -10 dB and -5 dB samples (p=0.1175), nor between the -10 dB and 0 dB samples (p=0.0782). However, the difference between -10 dB and +5 dB was significant (p=0.0055). At this highest level of noise, the AI system struggled to isolate the clinician’s and patient’s speech, leading to degraded accuracy and incomplete clinical summaries.

It is interesting to note that there is a strong correlation between the type of background noise and the number of errors introduced. The introduction of the toddler’s continuous, though unstructured chattering, caused a great number of errors.

At the other end of the spectrum, both the baby and construction noise caused far fewer errors. As there was no recognisable speech in these noises, the only mechanism for disruption was by literally drowning out the speech. However, the introduction of heavy rain (pink noise) at high volumes caused the greatest number of errors, suggesting that the frequency profile of certain background noises affects AI recognition differently. It also appears that intermittent background sounds, such as loud crashes, were better tolerated by the AI system compared to continuous noises.

### Impact of Informational Noise

The AI generated summary should capture only the important and relevant information. Examples of potentially confounding information are: (i) irrelevant medical-themed monologues by the patient, which should note dilute the clinician’s professional view, and not appear in the clinical summary. (ii) Casual conversations (for example, about the weather) should also be omitted from the clinical summary.

Medical and non-medical informational noise was added as either as a short snippet, or a longer rigmarole. Full details are in the supplementary information, but examples include:

- Patient very worried about unlikely cancer
- Patient very worried about unlikely chicken pox/shingles
- A protracted two-way discussion about tennis
- Patient complaining about the condition of the floor at their badminton club

When superfluous information was introduced into the audio file, either as a patient-derived-medical or conversational non-medical form, the clinical AI scribe was remarkably apt at reporting only the relevant details. Conversational pieces (for example, about a tennis club) were entirely left out, whilst irrelevant medical details (or speculative self-diagnoses) were either omitted or in some way tagged as being a patient opinion or concern.

Nevertheless, whilst this informational noise was usually not included (or was at least appropriately labelled), some errors still occurred. Figure 5 shows the absolute number of errors introduced into the summaries, whilst the relative error rate can be seen in figure S3 (supplementary information). The presence of a small number of inclusions is expected, as there may be new information to report (e.g. ‘patient worried about brain cancer’). However, the presence of omissions indicates that the proportion of medically relevant information captured has reduced. Throughout all experiments in this section, only a single hallucination was observed (see figure S3), which was merely a note to follow-up on a blood pressure check (the blood pressure check had been discussed, but the specific follow-up had not).

**Figure 5:**
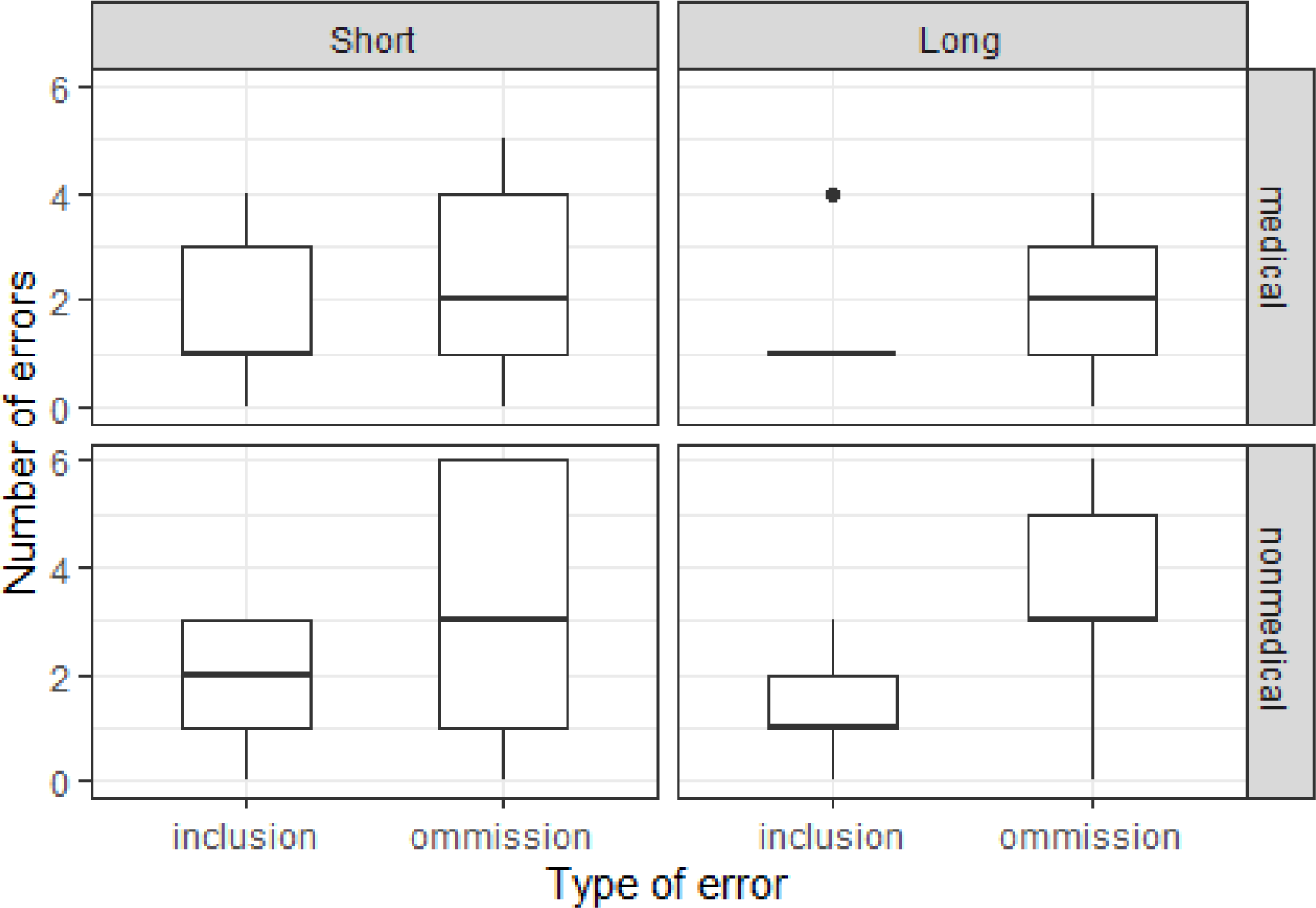
The number of errors (inclusions or omissions) introduced from the introduction of informational noise, from either non-medical conversations (bottom two plots) or patient-derived medical monologues (top two plots), for all medical scenarios combined. For each of these situations the quantity of informational noise is characterised as either short (left two plots) or long (right two plots). Transcripts of the inserted audio can be seen in the supplementary information.

Interestingly, there was no significant difference between the number of errors produced from irrelevant medical vs irrelevant non-medical information (p=0.6806, W=477 – Wilcoxon rank sum test with continuity correction). This suggests that the AI scribe is equally capable of rejecting both types of informational noise. Likewise, the difference between shorter and longer segments of informational noise was not significant when looking at omissions (p=0.846, W=463 – Wilcoxon rank sum test with continuity correction).

## Discussion

### Interpretation of Findings

The findings of this study show that this commercial Clinical AI Scribe can be highly sensitive to environmental conditions, particularly acoustic noise such as caused by microphone positioning or certain background noises. It should be noted however, that the errors produced add onto what is already a (typically very good, though) imperfect summary. We have observed that the commercial Clinical AI Scribe produces on average around 0.5 hallucinations and 1 omission per consultation under ideal audio conditions. (Further details on this will be presented in a future publication.)

The positive correlation between increased microphone distance and the rise in errors (specifically omissions) highlights the importance of microphone placement, set-up and testing in clinical settings. An unexpected discovery was that more expensive microphones did not necessarily provide better performance than cheaper microphones; and in fact, cheaper microphones could actually outperform them. The data was clear, however, that the integrated laptop microphone tested was particularly poor. It is suggested that a possible reason for this is a combination of the low sensitivity of the microphone (which could have been tuned to work optimally at typical laptop user distances), combined with the innate directionally of that microphone. It should also be noted that that the audio quality from different microphones in different laptops is, of course, highly variable [26].

The impact of background noise on accuracy emphasises the need for AI algorithms capable of filtering out environmental disturbances. It is evident that some types of noise, such as a baby crying, have minimal effect on the clinical summary generated. However, vocalisations, such as toddler chatter, caused notable degradation of the clinical summary. As the transcript does not record the toddler’s words, it appears that the errors are due to the Clinical AI Scribe struggling to isolate different voices when the toddler spoke over either the patient or the clinician. Additionally, and of particular interest, the pink noise generated by heavy rain also caused a particularly large error rate, even at lower (and more realistic) volumes. Due to its acoustic profile, pink noise is often used for sound masking [27], which may have contributed to the Clinical AI Scribe’s difficulty in distinguishing the speech.

Informational noise, in the form of either irrelevant casual conversation or patient-derived medical terms (for example, a speculative patient self-diagnosis) was treated comparatively well by the clinical AI scribe. Only a single hallucination was observed (with informational noise), though there were a number of omissions. As the source of the informational noise (medical or non-medical) did not have a statistically significant correlation with the number of omissions, it appears that (either through context, voice-identifying, or some other method) the clinical AI scribe is able to distinguish between the clinician and patient, and record their vocalisations with appropriate weighting. This is important because a clinical note should be weighted to a clinician’s thoughts, and not the patient’s [28].

Throughout the study, by far the most prevalent type of error were omissions. This demonstrates the conservative nature of this commercial Clinical AI Scribe, and suggests that the LLM has been designed, when presented with a difficult audio transcription, to err on the side of caution (producing omissions) rather than take a ‘best guess’ (which could produce hallucinations). It is generally accepted that that it is better to omit than to have incorrect information. Omissions observed included:

- Not recorded when the symptoms began.
- Not recording follow-up time.
- Not recording progression of symptoms.
- Not recording current medication.

Hallucinations were not a frequent type of error, though did occur, particularly with acoustic noise. Their appearance appeared random and unpredictable, with no hallucination repeated. Hallucinations observed included:

- Describes patient having tried epoxy, instead of ibuprofen.
- Called rash bumpy when it was explicitly stated as not.
- Recorded pain, when there was none.
- Reports unrelated pain in unrelated place.

With the exception of the Informational Noise section, it was interesting to note the presence of inclusions (accurate points of information present in the new summary, but not the original summary). It was anticipated that the addition of either type of noise would cause a decrease in the number of points of information, but instead, in several cases, the clinical AI scribe noted additional truthful points which it had previously ignored. For example, the addition of a conversation in which the clinician and patient are socially friends and discuss babysitting, mother-in-laws, and arranging a playdate caused the AI to accurately report that the patient is a smoker in the clinical summary (the smoking discussion was temporally far removed from the inserted casual conversation). Whilst this information was accurate (it was mentioned in the original audio), it was not reported in the original summary. The mechanism behind this remains unclear, though it demonstrates the LLMs unpredictable sensitivity to user input.

### Implications for Clinical Practice

Given the rapidly increasing international uptake of AI-powered documentation tools in primary care (and elsewhere in clinical workflows), it is essential to ensure the accuracy and reliability of these systems. These findings underscore several key considerations for the integration of AI scribe technology into primary care settings.

Firstly, microphone placement should be optimised within the consultation environment, with recommendations favouring a dedicated microphone positioned within one metre of the clinician, to reduce omission errors. In particular, integrated laptop microphones should be avoided. It is possible that lapel microphones could offer an improvement, although this could impact user acceptance. Additionally, an additional microphone located at an examination couch (which could be upwards of 2 metres away [29]) may be required to ensure all conversation is captured. Secondly, whilst the AI clinical scribe was generally able to accommodate low levels of background noise, interference from other vocalisations (such as toddlers) or pink noise (such as heavy rain) should be carefully minimised. This is especially important when the noise volume is equal or greater than the volume of the speech. Thirdly, when non-medically relevant social conversations are present in a consultation, additional care should be taken in confirming the generated summary, to ensure that no omissions have occurred.

The issue of hallucinations and omissions in AI-generated clinical summaries reinforces the fact that (as required by all commercial providers) clinician oversight remains a necessary safeguard. While AI has the potential to streamline documentation [1], its current limitations indicate that all generated summaries should undergo manual review before being integrated into patient records. Clinicians must be trained to identify potential AI errors and to verify the accuracy of recorded information. It must be emphasised that user overconfidence in the accuracy of the AI-generated summary should be guarded against, as errors occur regularly enough to be problematic, but infrequently enough to risk complacency. And whilst the effect of such an event would be variable, it is quite possible that delays in diagnosis, impacts on treatment, or even a fatal consequence may occur.

### Future Directions for Research

While this study provides significant insights into the challenges associated with Clinical AI Scribes, further research is needed to understand and enhance the robustness and usability of these systems. It is important to acknowledge that these tools are undergoing swift advancements, and considerable enhancements in performance are anticipated in the near future.

One important area for future study is the development of AI models trained on datasets that incorporate a greater variety of real-world clinical noise conditions. There are currently very few publicly available databases which also include the audio file, e.g. [18]. Additionally, investigations into multimodal AI scribe approaches, which integrate audio, text, and contextual patient data, may yield improvements in accuracy and relevance.

Another avenue of exploration is the impact of AI scribe on clinician workload and efficiency. While AI scribes offer the potential to reduce administrative burden, it remains to be seen whether their implementation leads to measurable improvements in workflow and patient outcomes [4,30]. Longitudinal studies assessing the long-term effectiveness of AI-assisted documentation, including its influence on consultation dynamics and medico-legal considerations, will be essential for guiding future AI adoption in healthcare.

## Conclusion

AI-powered clinical scribes have great potential to improve clinical documentation and alleviate administrative burdens for healthcare professionals [1,2]. This study highlights the challenges associated with the use of Clinical AI scribes, particularly its sensitivity to microphone quality/positioning and environmental noise.

The findings reinforce the importance of microphone placement, with close-proximity, dedicated microphones yielding the most reliable results. Increased distance and the use of integrated microphones were associated with higher omission rates, suggesting that optimised audio capture strategies should be a priority in clinical settings. Background noise, particularly comprehensible speech (for example toddler chatter) and heavy rain (pink noise), also significantly impacted accuracy. Whilst both non-medical and medical informational noise (for example a conversation about tennis or speculative patient self-diagnosis), did contribute to an increase in errors, they were far less pronounced than those caused by acoustic noise.

Ultimately, clinician oversight remains a key factor in ensuring accuracy of both the summary and the EHR. While AI has the potential to streamline documentation, human validation is necessary to correct potential errors. Structured review workflows will be crucial to maintaining data integrity and supporting safe clinical decision-making.

## Supporting information

Protocols/Method

## Acknowledgements

The authors thank NHS England South West for the funds used to establish the CoDE at UWE Bristol, and the participants of the Citizens Reference Group for their contributions.

## Contributions

JMC, ST, JK, and RL conceived and designed the study. TCD, JL, KLR, and TC were involved in the acquisition of data. TCD, JL, TC, and RL analysed the data. TCD, TC, and JMC drafted the manuscript. All authors approved the final version of the manuscript. The corresponding author attests that all listed authors meet authorship criteria and that no others meeting the criteria have been omitted.

## Ethical Approval

Ethical approval for the work undertaken was granted by the University of the West of England Ethics committee – No: CHSS.24.05.199.

## Data Availability Statement

Outputs from the commercial Clinical AI Scribe, along with transcripts of the original audio files, will be made available upon reasonable request to the corresponding author.

## Transparency Statement

The corresponding author affirms that the manuscript is an honest, accurate, and transparent account of the study being reported; that no important aspects of the study have been omitted; and that any discrepancies from the study as planned have been explained.

