## Supplementary material for "The Impact of Acoustic and Informational Noise on AI-Generated Clinical Summaries": Protocols/Method

(b) NHS England South West, South West House, Blackbrook Ave, Taunton, TA1 2PX, UK

### Protocols

#### Recording of Simulated Medical Consultations

Simulated consultations were performed by three actors (in role rotation as either clinician or patient), focusing on one of five common primary care scenarios: memory loss, diarrhoea, headaches, skin rash, or prostate symptoms. The consultations were recorded using 2 video cameras (one directed towards the front of each actor) and a dedicated microphone. The audio recordings were generated as wav format files. These recorded simulated consultations provided the basis for the following experiments, where the acoustic signal-to-noise at the input to the automatic speech recognition module (ASR) (i.e. how clear the audio is) and the informational signal-to-noise (i.e. ratio of clinically significant to irrelevant information) are varied. All of the experiments to investigate the performance of the LLM based clinical AI scribe were conducted between August 2024 and January 2025.

#### Effect of Microphone Placement (Acoustic Noise)

In order to determine the effect of the selected microphone on the acoustic signal-to-noise (and the corresponding impact on the semantic signal-to-noise), four different microphones were tested. Audio files of five different medical consultations were played through dedicated speakers (Logitech S-150) to the four microphones. The audio files used were stereo, such that the clinician spoke only out of one channel, and the patient out of the other. The speakers were positioned 1.0 m apart, in such a way as to emulate the typical positions of a clinician and patient during a consultation.

The microphones used in the study were: (a) Konftel Ego (omni-directional conference mic, RRP £120), (b) Fifine K668 (metal bodied 'traditional' mic, RRP £25), (c) Neat Skyline (upright mic, RRP £40), and (d) the integrated microphone of an HP Elitebook 840 G3 laptop. All four microphones were tested at 0.5 m, 2.0 m, and 4.5 m from the midpoint of the speakers. A direct line of sight was maintained between the microphone and the speakers, apart from at 4.5 m, where there was additionally a small examination screen obstructing. The automatic gain control was disabled for all experiments.

The stereo audio files were played (5), recorded through each of the microphones (4) at each position (3). These 60 audio files were then presented to the commercial clinical AI scribe, from which 60 clinical summaries were produced. The default settings for the clinical AI scribe were used, along with the Subjective, Objective, Assessment, Plan (SOAP) template.

To analyse these clinical summaries, the original audio recordings were also presented to the clinical AI scribe, generating a clinical summary in the same manner as above – this clinical summary was considered the 'ground truth' for its respective consultation. This is acceptable because this study was concerned with variation in the output when the input is modified, not the core abilities of the commercial clinical AI scribe. Each of the 60 clinical summaries was then manually compared to its corresponding 'ground truth', with the total 'points' of information present, the corresponding number of omissions, and any hallucinations or inclusions counted.

**Addition of Background Noise (Acoustic Noise)**

In order to investigate the effect of background noise in a controlled manner, audio recordings of five different medical consultations were modified to include a specific amount of background noise. Four different background noises, stored as lossless audio files, were chosen (for source details, see supplementary information), to represent typical auditory distractions: baby crying, construction, heavy rain, and toddler chatter. Using the Python Pydub library (https://pypi.org/project/pydub/), each of these different sounds (4) were combined with each of the different consultations (5), at four different relative volumes: -10 dB, -5 dB, 0 dB, and +5 dB.  The background noise files were not normalised against the consultation volumes, but were of similar loudness. The 80 generated audio files were then presented to the clinical AI scribe using the previously mentioned settings, generating 80 clinical summaries.

Analysis of these 80 clinical summaries was performed in a similar manner to above: each was manually compared to the clinical summary generated using the original unmodified recording, and any omissions, hallucinations, and inclusions noted.

**Addition of Irrelevant Medical/Non-Medical Information (Informational Noise)**

A summary of a medical consultation generated by a clinical AI should ideally only contain the relevant medical details. In order to examine this, the audio recordings of five medical consultations were modified using Audacity (https://www.audacityteam.org/) to include additional conversations between the clinician and the patient. Into the middle of each of the five original recordings was spliced an additional recording (by the same actors), which would contain either a short or long irrelevant medical or non-medical conversation, thereby generating 20 new audio files. These were then presented to the clinical AI scribe using the previously described settings, generating 20 clinical summaries.

Examples of the types of conversations inserted include: (i) discussing meeting up with mutual friends, (ii) discussion of the local tennis club, and (iii) unfounded concerns about severe illness. Full scripts can be found in the supporting information.

Analysis of the 20 generated clinical summaries was performed in a similar manner to above: each was manually compared to the clinical summary generated using the original unmodified recording, and any omissions, hallucinations, and inclusions noted.

**Statistical Analysis**

Statistical analyses, using nonparametric tests, were conducted to evaluate the significance of observed variations. Statistical analysis was conducted using R (R Core Team (2024). _R: A Language and Environment for Statistical Computing_. R Foundation for Statistical Computing, Vienna, Austria. https://www.R-project.org/) in R Studio (Posit team (2024). RStudio: Integrated Development Environment for R. Posit Software, PBC, Boston, MA. URL http://www.posit.co/). Nonparametric analysis was used to compare number of errors and error rates, employing the Exact two-sample Kolmogorov-Smirnov test and Mann-Whitney U 2 tailed test and Wilcoxon rank sum test with continuity correction. Significance is taken as $p < 0.05$.
