## Supplementary Information for "The Impact of Acoustic and Informational Noise on AI-Generated Clinical Summaries"

(b) NHS England South West, South West House, Blackbrook Ave, Taunton, TA1 2PX, UK

### Supplementary Information

#### Figures

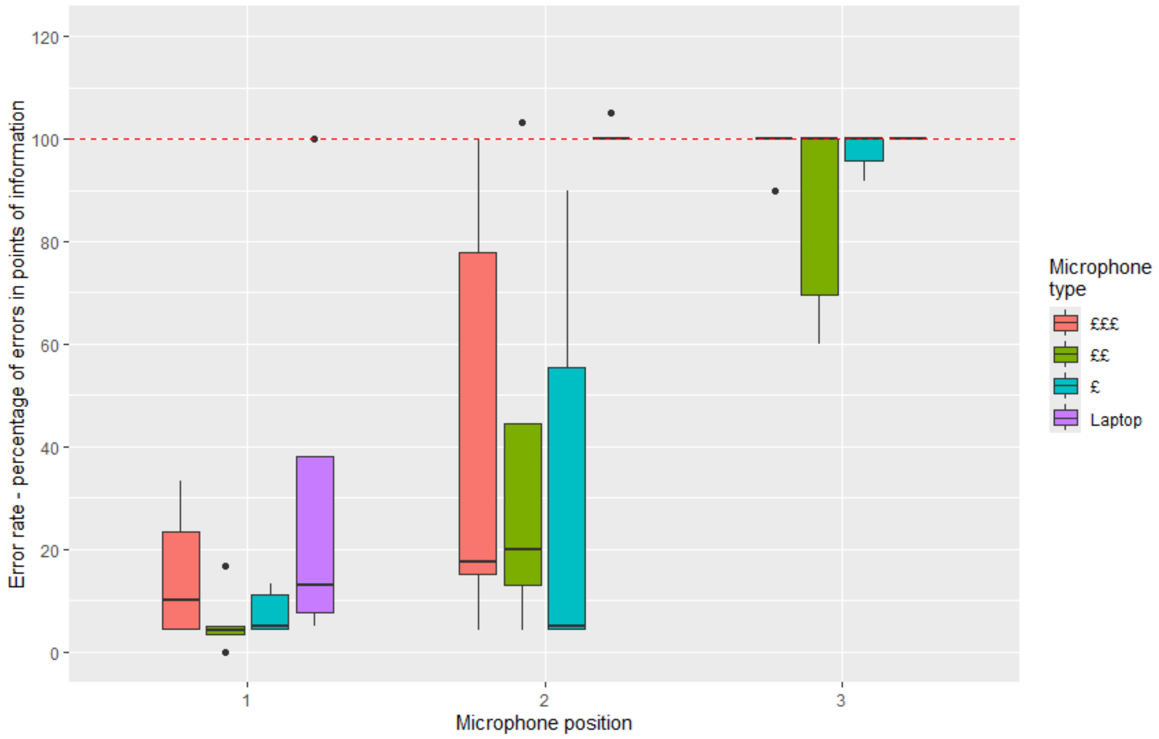


*S1 – Figure to show the percentage reduction in points of information, according to cost of the microphone. The red line indicates 100%, i.e. complete loss of all information. 0% refers to the original summary, which is assumed to be accurate. Microphone positions: d= (1) 0.5m, (2) 2.0m, and (3) 4.5m.*

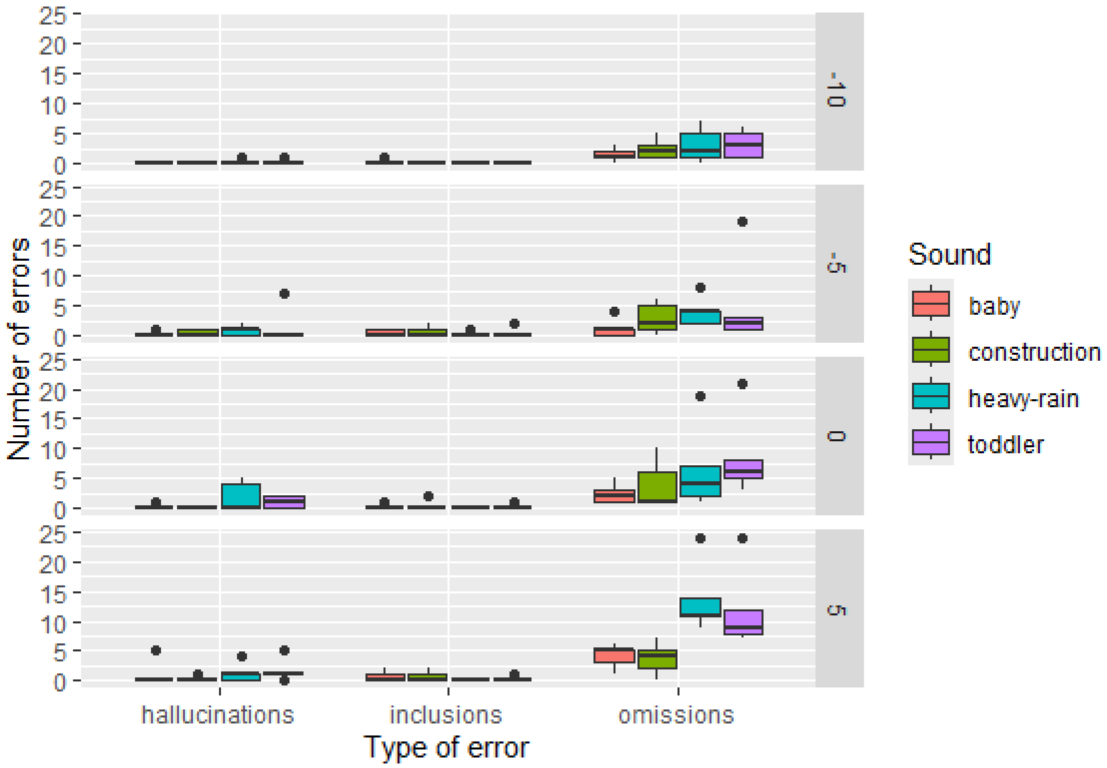


*Figure S2. The number of the different types of errors induced by background noise at varying levels with respect to the consultation: -10, -5, 0 and +5 dB.*

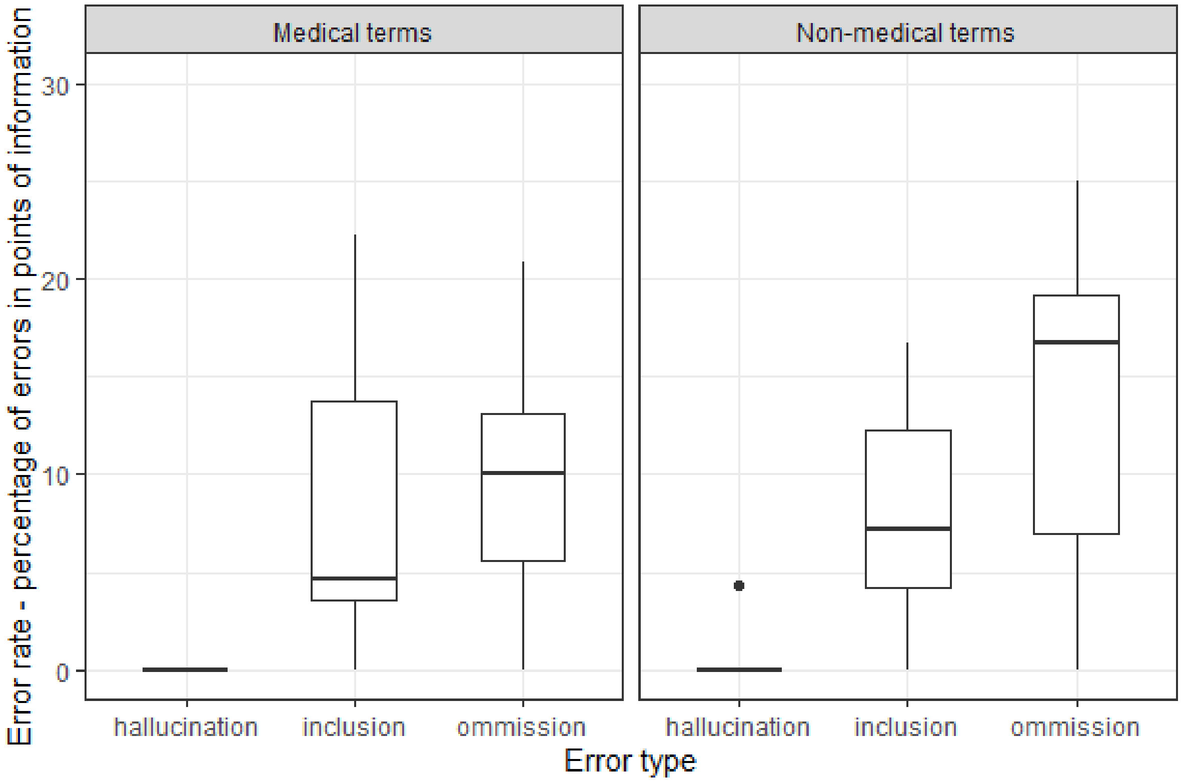


*Figure S3: The relative error rate (as a percentage) caused by the introduction of informational noise, from either speculative self-diagnosis or excessive casual conversation in a consultation. Amalgamation of short and long for all consultation scenarios.*

### Background Noise

All background noise files were sourced from freesound.org.

*Construction Noise*: 190611_MiniExcavator.flac by tillt -- https://freesound.org/s/474442/ -- License: Creative Commons Attribution 4.0 International

*Baby*: 585111__nome_diva__baby Baby.wav by nome_diva -- https://freesound.org/s/585111/ -- License: Creative Commons CC0 1.0 Universal

*Toddler*: 416704__funwithsound__baby-gibberish-and-words-full.wav Baby Gibberish and Words - Full.wav by FunWithSound -- https://freesound.org/s/416704/ -- License: Creative Commons CC0 1.0 Universal

*Rain*: 353237__themysteriousmrx__1_hour_of_hurricaane_hermine.ogg 1_hour_of_hurricaane_hermine.ogg by themysteriousmrx -- https://freesound.org/s/353237/ -- License: Creative Commons CC0 1.0 Universal

### Informational Noise Scripts – Non-medical

**Scenario: Skin rash**

**Low level of additional information**

P: Good to see you, it's been a few weeks, how's Gill doing.

GP: Yes, very well she's started back at work and enjoying it.

P: Is the child care working out as I know that was something you were worried about.

GP: Sara has settled into the nursery, she gets a bit tired and grumpy towards the end of the week but nothing too bad.  Anyhow why have you come to see me today?

**High level of additional information**

P: Good to see you, it's been a few weeks, how's Gill doing.

GP: Yes, very well she's started back at work and enjoying it, although she still feels guilty about not being at home for the twins.

P: Is the child care working out as I know that was something you were worried about.

GP: Sara has settled into the nursery, she gets a bit tired and grumpy towards the end of the week but nothing too bad.  Maisy loves it, she's the outgoing one. Getting out in the morning can be a bit hectic but having Gill's Mum close by helps, I never thought I'd say that!

P: I remember when Julia went back to work we had the same issues, with us it was arranging the pick up from nursery, we both had to be really disciplined with meetings at the end of the day so we could get away on time.  We should arrange a date so the four of us can catch up. It's our turn to host.

GP: If you send a few dates over we'll get Gill's Mum to babysit, we'll just have to check when she's free.  Anyhow what can I do for you today?

**Scenario: Headache**

**Low level of additional information**

P: Hello Dr, didn't I see you at the tennis club last night?

GP: Yes, my evening surgery times have changed so I can go to the Wednesday night club session now.

P: I've started going again on Wednesday, just trying to get a bit of fitness back.

GP: Well I may see you next week, what can I do for you today?

**High level of additional information**

P: Hello Dr, didn't I see you at the tennis club last night?

GP: Yes, my evening surgery times have changed so I can go to the Wednesday night club session now.

P: I've started going again on Wednesday, just trying to get a bit of fitness back.

GP: I haven't seen you down at the club for a number of weeks, why's that.

P: A combination of things, family events and I was away with work a lot, we started a new outlet up in Chester.

GP: Explains why you've not been playing?

P: Yes. I'm aiming to be fit for the summer league. Are you going to enter?

GP: Not sure, Friday nights are busy and I'm not quite your standard, so I may just fill in when required. Anyhow what can I do for you today?

**Scenario: Memory Loss**

**Low level of additional information**

GP: Good afternoon Mrs ******** how can I help today?

P: Nothing to do with why I'm limping, I did that last night playing badminton.

GP: Is it painful?

P: No it's just a slight tweak of the muscle, nothing major I do it all the time, I'll recover in a couple of weeks. The issue is that the floor at the leisure centre is slippery, they polish it too much a number of players hurt themselves over stretching. We have asked them not to buff the floor as often but they won't listen.

**High level of additional information**

GP: Good afternoon Mrs ******** how can I help today?

P: Nothing to do with why I'm limping, I did that last night playing badminton. Just a tweak of the calf muscle. I've seen a physio about it before so I know what exercises I need to do.  I also know I need to rest up for a couple of weeks whilst it heals.  So not a problem really.

GP: Is it painful?

P: I'll recover in a couple of weeks.  The issue is that the floor at the leisure centre is slippery, they polish it too much a number of players hurt themselves over stretching. We have asked them not to buff the floor as often but they won't listen.  Although I played on we lost the game as my movement was restricted. It's a pity as we won the first game easily and we've beaten that pair before on a number of occasions.  It gets frustrating.  Last week the same thing happened to the visiting team.  It's disappointing you want a full evening out playing not sitting on the side lines watching.  It's even worse for my partner he's very competitive and didn't like losing to that pair, particularly since they were from his old club.

GP: So, if it's not the leg then what's the problem today?

**Scenario: Diarrhoea**

**Low level of additional information**

GP:  Good morning Mr ******* how can I help you today?

P: I'm not sure I don't think there's anything wrong.

GP: (Pause)

P: My partner suggested that I came to see you.

GP: What was your partner concerned about?

P: I don't think it's anything really, she can fuss over things but........

**High level of additional information**

GP:  Good morning Mr ******* how can I help you today?

P: I'm not sure I don't think there's anything wrong.

GP: (Pause)

P: My partner suggested that I came to see you.

GP: What was your partner concerned about?

P: I don't think it's anything really, she can fuss over things, once she gets something in her head it takes some shifting.

GP: If you can let me know what is worrying her then we can hopefully put her mind to rest.

P: Well it's personal and I don't like to talk about it.

GP: I can assure you whatever you say will remain between the two of us, it's my job to help with personal issues.

P Well it's embarrassing but………

**Scenario: Prostrate symptoms**

**Low level of additional information**

GP: Good morning Mr ******* how can I help you today?

P: I'm not sure I don't think there's anything wrong.

GP: (Pause)

P: My wife suggested that I came to see you.

GP: What was your wife concerned about?

P: I don't think it's anything really, she can fuss over things but……..

**High level of additional information**

GP: Good morning Mr ******* how can I help you today?

P: I'm not sure I don't think there's anything wrong.

GP: (Pause)

P: My wife suggested that I came to see you.

GP: What was your wife concerned about?

P: I don't think it's anything really, she can fuss over things, once she gets something in her head it takes some shifting.

GP: If you can let me know what is worrying her then we can hopefully put her mind to rest.

P: Well it's personal and I don't like to talk about it.

GP: I can assure you whatever you say will remain between the two of us, it's my job to help with personal issues.

P: Well it's embarrassing but………

### Informational Noise Scripts – Medical

**Scenario: Skin rash**

**Low level of additional information**

P: One of my friends has shingles it looks similar, and shingles has spread across his arms and back so I thought that shingles was a probable diagnosis.  There's a lot of it going around at present.

**High level of additional information**

P: One of my friends has shingles it looks similar, and shingles has spread across his arms and back so I thought that shingles was a probable diagnosis.  There's a lot of it going around at present.

P: I did have chickenpox when I was a child and I know that the virus from chickenpox stays in the systems for the rest of your life and can cause shingles. As I said when it spread onto my back I thought I should come and see you.

**Scenario: Headache**

**Low level of additional information**

P: My uncle developed double vision so it was something I was aware of and kept an eye on.  I thought I may have a brain tumour or something.  I know it sounds dramatic but it does happen, you read things in the press about people even younger than me with brain tumours.

**High level of additional information**

P: My uncle developed double vision so it was something I was aware of and kept an eye on.  I thought I may have a brain tumour or something.  I know it sounds dramatic but it does happen, you read things in the press about people even younger than me with brain tumours.

P: I feel that as the headaches have gone on for so long I can't rule out a brain tumour. Things start that way and although I don't have double vision at the moment it could develop.  Sorry I feel as though I'm being dramatic.

**Scenario: Memory Loss**

**Low level of additional information**

P: There's been a lot in the news about people with dementia and other things like Alzheimer's and Parkinson's disease. You can't help but be worried.

**High level of additional information**

P: There's been a lot in the news about people with dementia and other things like Alzheimer's and Parkinson's disease. You can't help but be worried.

P: I used to be really organised and on the ball. I just worry that I'm losing part of myself to some disease that is slowly eating away at my brain. I know that you can get things such as early onset Alzheimer's as a form of dementia. Although I'm relatively young I'm not immune from these things and I also know that although there's no cure for dementia there are drugs that can help and delay the progression of diseases such as Alzheimer's or Parkinson's.

**Scenario: Diarrhoea**

**Low level of additional information**

P: I am worried about cancer I have googled things and I known that it could be cancer related. I think that bowel cancer is a possibility. I also know that bowel cancer is hard to treat and it would mean operations and chemotherapy.

**High level of additional information**

P: I am worried about cancer I have googled things and I known that it could be cancer related. I think that bowel cancer is a possibility. I also know that bowel cancer is hard to treat and it would mean operations and chemotherapy.

P: Reading around I know that it could also be other cancers. I haven't been scanned for polyps in my colon and this could be related to that. Again, that could be a cancer of at best some form of inflammatory bowel syndrome. All of it could be pretty horrific, all of these cancers involve a lot of invasive treatments and post operation the chemo and radio therapy are unpleasant.

**Scenario: Prostrate symptoms**

**Low level of additional information**

P: However, I'm hoping it could be something as simple as a urinary tract infection a UTI. I've had a few UTIs in the past and they have been cleared up with antibiotics. I haven't had one for a couple of years so I'm almost due a UTI.

**High level of additional information**

P: However, I'm hoping it could be something as simple as a urinary tract infection a UTI. I've had a few UTIs in the past and they have been cleared up with antibiotics. I haven't had one for a couple of years so I'm almost due a UTI.

P: When I have a UTI I do go to the toilet a lot, and it can be painful and I can have trouble going so it is a bit similar, so I can't rule a UTI out although there is still the obvious red flag.
